# The increase in the risk of severity and fatality rate of covid-19 in southern Brazil after the emergence of the Variant of Concern (VOC) SARS-CoV-2 P.1 was greater among young adults without pre-existing risk conditions

**DOI:** 10.1101/2021.04.13.21255281

**Authors:** André Ricardo Ribas Freitas, Daniele Rocha Queiróz Lemos, Otto Albuquerque Beckedorff, Luciano Pamplona de Góes Cavalcanti, Andre M Siqueira, Regiane Cristina Santos de Mello, Eliana N C Barros

## Abstract

**Background:** The SARS-CoV-2 P.1 variant has been considered as “variant of concern (VOC)” since the end of 2020 when it was firstly identified in the Brazilian state of Amazonas and from there spread to other regions of Brazil. This variant was associated with an increase in transmissibility and worsening of the epidemiological situation in the places where it was detected. The aim of this study was to analyze the severity profile of covid-19 cases in the Rio Grande do Sul state, southern region of Brazil, before and after the emergence of the P.1 variant, considering also the context of the hospitals overload and the collapse of health services.

**Methods:** We analyzed data from the Influenza Epidemiological Surveillance Information System, SIVEP-Gripe (Sistema de Informação de Vigilância Epidemiológica da Gripe) and compare two epidemiological periods: the “first wave” comprised by cases occurred during November and December 2020 (EW 45 to 53) and the “second wave” with cases occurred in February 2021 (EW 5 to 8), considering that in this month there was a predominance of the new variant P.1. We calculated the proportion of severe forms among the total cases of covid-19, the case fatality rates (CFR) and hospital case fatality rate (hCFR) over both waves time set using the date of onset of symptoms as a reference. We analyzed separately the patients without pre-existing conditions of risk, by age and sex. For comparison between periods, we calculated the Risk Ratio (RR) with their respective 95% confidence intervals and the p-values.

**Findings:** We observed that in the second wave there were an increase in the proportion of severe cases and covid-19 deaths among younger age groups and patients without pre-existing conditions of risk. The proportion of people under the age of 60 among the cases that evolved to death raised from 18% (670 deaths) in November and December (1^st^ wave) to 28% (1370 deaths) in February (2^nd^ wave). A higher proportion of patients without pre-existing risk conditions was also observed among those who evolved to death due to covid-19 in the second wave (22%, 1,077 deaths) than in the first one (13%, 489 deaths). The CFR for covid-19 increased overall and in different age groups, in both sexes. The increase occurred in a greatest intensity in the population between 20 and 59 years old and among patients without pre-existing risk conditions. Female 20 to 39 years old, with no pre-existing risk conditions, were at risk of death 5.65 times higher in February (95%CI = 2.9 - 11.03; p <0.0001) and in the age group of 40 and 59 years old, this risk was 7.7 times higher (95%CI = 5.01-11.83; p <0.0001) comparing with November-December.

**Interpretation:** Our findings showed an increase in the proportion of young people and people without previous illnesses among severe cases and deaths in the state of RS after the identification of the local transmission of variant P.1 in the state. There was also an increase in the proportion of severe cases and in the CFR, in almost all subgroups analyzed, this increase was heterogeneous in different age groups and sex. As far as we know, these are the first evidences that the P.1 variant can disproportionately increase the risk of severity and deaths among population without pre-existing diseases, suggesting related changes in pathogenicity and virulence profiles. New studies still need to be done to confirm and deepen these findings.

## Introduction

Since the beginning of 2021, international authorities have shown great concern with the P.1 variant of SARS-CoV-2, also known as 20J / 501Y.V3 or Variant of Concern 202101/02 (VOC-202101/02). This variant probably emerged between late November and early December 2020 in Amazonas, northern Brazil, with rapid dissemination to other regions and other countries[1]. The introduction of this variant was temporally associated with increased transmissibility of covid-19 causing a critical epidemiological scenario in different places where it was detected as the Amazonas, in the north region, and the Paraná and the Rio Grande do Sul (RS) states, in the south of the country[2–5]. In the Amazonas state, a collapse of the health system was observed, which made it difficult to assess the real impact of the P.1 variant on the lethality by covid-19, which may have increased due to the overload of the health system and not exclusively by the intrinsic characteristics of the variant[2]. The emergence of variant P.1 in the RS state was confirmed by the virus identification from a patient with no travel history and presenting symptoms beginning on January 29 (EW 4/2021)[6]. In the following weeks there was a sudden increase in cases of covid-19 in several regions of the state, simultaneously. Virological surveillance data showed that in February, the variant P.1 corresponded to about 70% of the viruses sequenced in the RS state. [5,7]

The exponential increase in the hospitalization rate observed in the RS state could be explained, on one hand, by the P.1 circulation as this variant has being 2.6 times more transmissible (95% Confidence Interval (CI): 2.4–2.8) than the previous one [8]. However, on other hand, studies have suggested that the P.1 variant can also lead to more severe conditions, which would increase the need for hospitalization and contribute to the increase observed in hospitalization rates. In order to provide more evidences related to the change in the hospitalization and deaths patterns among covid-19 cases in the RS state, we performed an epidemiological analysis describing and comparing the severity and mortality profile of covid-19 cases in the RS state, considering two periods before and after the emergence of variant P.1.

## Methodology

An observational, retrospective epidemiological study of covid-19 cases reported in the National Influenza Epidemiological Surveillance Information System (SIVEP-Gripe) of the Ministry of Health of Brazil was performed.

### Database and setting

The Rio Grande do Sul state has 11,422,973 inhabitants, it is the 5^th^ largest state in population size in the country, corresponding to 5.4% of the Brazilian population. Life expectancy at birth is 78.0 years and the HDI is 0.787[9]. The Unified Health System (SUS), the public health system in Brazil, is responsible for 72% of the 3,411 ICU beds available in the state[7].

The notification of suspected and confirmed cases of covid-19 is mandatory in Brazil, both in public and private health services. Suspected cases of covid-19 can be confirmed by laboratory, clinical, clinical-epidemiological or clinical-radiological criteria, according to the Brazilian guidelines (supplements) that follow the recommendations of the World Health Organization. In the SIVEP-Gripe system, cases can be classified as influenza syndrome (influenza-like illness (ILI)) and severe cases (severe acute respiratory infections (SARI)). SARI cases are confirmed cases of covid-19 requiring hospitalization associated with any of the following signs and symptoms: dyspnea, difficulty breathing, O2 saturation below 95% in ambient air or cyanosis. In children, in addition to the previous items, the following nasal wing beats, intercostal circulation, dehydration and lack of appetite are included.

We accessed information on demographic data, self-reported ethnicity, presence of pre-existing risk conditions (supplements), data on the onset of symptoms, hospitalization and hospital outcome, as well as data on the occupation of ward beds and intensive care unit (ICU) beds. The data were made available by the Secretariat of Health of the Rio Grande do Sul state and by the Ministry of Health of Brazil through open access online platforms with anonymous data. The period of analysis was from the epidemiological week 17/2020 (started on April 19^th^ 2020) to the epidemiological week 11/2021 (ended on March 20^th^ 2021). The data were exported for analysis on April 2^nd^ 2021. [7,10]

### Data analysis

We defined two epidemiological periods for the analysis: the “first wave” comprised by cases occurred during November and December 2020 (EW 45 to 53) and the “second wave” with cases occurred in February 2021 (EW 5 to 8), considering that in this month there was a predominance of the new variant P.1. The month of March 2021 was not included in the second wave due to the fact that, at the beginning of this month, the health system reached 100% of the occupancy of ICU beds, which may affect the risk of death.

We calculated the proportion of severe forms among the total cases of covid-19, the case fatality rates (CFR) and hospital case fatality rate (hCFR) over both waves time set using the date of onset of symptoms as a reference. The hCFR was calculated dividing the number of deaths by the total number of patients who had already been classified by hospital discharged or death[11].

We calculated the proportion of severe forms (SARI) between the total number of covid-19 cases and the case fatality rate (CFR), in the first and second waves. We analyzed separately the patients without pre-existing conditions of risk, by age and sex. For comparison between these periods, we calculated the Risk Ratio (RR) with their respective 95% confidence intervals and the p-values.

Additionally, the bed occupancy rate in the ICU was used as an indicator of the capacity of the local health system because it is the level of assistance required for the life risk patients management and it was considered by the health sector in the RS state as the principal indicator to signalize the exhaustion of the hospital capacity during the pandemic.

The data were analyzed using the STATA 16 software and followed the recommendations of the **RE**porting of studies **C**onducted using **O**bservational **R**outinely-collected **D**ata (**RECORD**) guidelines[12], attached checklist. This study did not require approval from any research ethics committee as all data were anonymous and obtained from open and public open source databases.

## Results

In the state of Rio Grande do Sul, 230,986 cases of covid-19 were confirmed in the first wave and 150,942 cases in the second wave, while the number of severe cases was 11,951 and 13,128, respectively (table 1). Mortality was also higher in the second wave (4,859 deaths) when compared to the first wave (3,809 deaths). The proportion of cases of covid-19 by age group did not change between the first and second waves, however, the proportion of people under 60 years of age among severe cases increased from 39% in the first wave to 47 %, in the second wave. Also, the proportion of people under the age of 60 among the cases that evolved to death raised from 18% (670 deaths) in November and December (1^st^ wave) to 28% (1370 deaths) in February (2^nd^ wave) (data summarized from the Table 1).

**Table 1:** Demographic, underlying conditions and diagnostic criteria of cases, severe cases and deaths due to covid-19 during the first (Nov-Dec/2020 and second waves (Feb/2021) in the Rio Grande do Sul state, Brazil.

|  | Covid-19 (all cases) |  |  |  | Severe |  |  |  | Deaths |  |  |  |
| --- | --- | --- | --- | --- | --- | --- | --- | --- | --- | --- | --- | --- |
|  | Nov-Dec/2020 |  | Feb/2021 |  | Nov-Dec/2020 |  | Feb/2021 |  | Nov-Dec/2020 |  | Feb/2021 |  |
| N | 230996 |  | 150952 |  | 11961 |  | 13138 |  | 3809 |  | 4859 |  |
| Age group |  |  |  |  |  |  |  |  |  |  |  |  |
| 0-19 ys | 20548 | 9% | 14545 | 10% | 116 | 1% | 88 | 1% | 4 | 0% | 10 | 0% |
| 20-39 ys | 96716 | 42% | 61910 | 41% | 1038 | 9% | 1615 | 12% | 100 | 3% | 241 | 5% |
| 40 - 59 ys | 75456 | 33% | 49107 | 33% | 3449 | 29% | 4473 | 34% | 566 | 15% | 1119 | 23% |
| 60 - 79 ys | 33092 | 14% | 22114 | 15% | 5395 | 45% | 5454 | 42% | 2017 | 53% | 2496 | 51% |
| 80 and more ys | 5184 | 2% | 3276 | 2% | 1963 | 16% | 1508 | 11% | 1122 | 29% | 993 | 20% |
| Sex |  |  |  |  |  |  |  |  |  |  |  |  |
| Female | 124882 | 54% | 80912 | 54% | 5414 | 45% | 5955 | 45% | 1699 | 45% | 2267 | 47% |
| Ethnicity |  |  |  |  |  |  |  |  |  |  |  |  |
| White | 166105 | 72% | 110900 | 73% | 8835 | 74% | 9116 | 69% | 1754 | 46% | 1960 | 40% |
| Mixed ethnicity | 8100 | 4% | 6125 | 4% | 396 | 3% | 343 | 3% | 86 | 2% | 80 | 2% |
| Black | 7857 | 3% | 4860 | 3% | 410 | 3% | 367 | 3% | 76 | 2% | 82 | 2% |
| Asian | 1553 | 1% | 408 | 0% | 21 | 0% | 24 | 0% | 5 | 0% | 2 | 0% |
| Indigenous | 321 | 0% | 95 | 0% | 31 | 0% | 14 | 0% | 6 | 0% | 2 | 0% |
| N. D. | 47050 | 20% | 28554 | 19% | 2258 | 19% | 3264 | 25% | 1882 | 49% | 2733 | 56% |
| Diagnostic criteria |  |  |  |  |  |  |  |  |  |  |  |  |
| RT-PCR | 146808 | 64% | 82043 | 54% | 9809 | 82% | 9221 | 70% | 3339 | 88% | 3595 | 74% |
| Immunochromatographic test | 75806 | 33% | 65666 | 44% | 1354 | 11% | 3167 | 24% | 329 | 9% | 1081 | 22% |
| Another test | 5960 | 3% | 1954 | 1% | 133 | 1% | 121 | 1% | 34 | 1% | 31 | 1% |
| Clinical criteria | 1430 | 1% | 682 | 0% | 51 | 0% | 127 | 1% | 1 | 0% | 11 | 0% |
| Clinical and image criteria | 640 | 0% | 444 | 0% | 581 | 5% | 444 | 3% | 97 | 3% | 133 | 3% |
| Clinical-epidemiological criteria | 339 | 0% | 132 | 0% | 20 | 0% | 27 | 0% | 8 | 0% | 5 | 0% |
| N. D. | 3 | 0% | 21 | 0% | 3 | 0% | 21 | 0% | 1 | 0% | 3 | 0% |
| Underlying conditions |  |  |  |  |  |  |  |  |  |  |  |  |
| No | 201320 | 87% | 130612 | 87% | 3021 | 25% | 4324 | 33% | 489 | 13% | 1077 | 22% |

There was no change in the proportion of women among severe covid-19 cases or among deaths between the two analyzed periods and the same profile was observed in the proportion of covid-19 cases without pre-existing risk conditions keeping around 87% in the two waves (87%). However, the proportion of patients without pre-existing risk conditions among severe cases was higher in the second wave (33%, 4,324 severe cases) than in the first wave (25%, 3,021 severe cases) (Table 1). A higher proportion of patients without pre-existing risk conditions was also observed among those who evolved to death due to covid-19 in the second wave (22%, 1,077 deaths) than in the first one (13%, 489 deaths) (Table 1).

After the emergence of the variant P.1 (week 4/2021) in the RS state a rapid increase in the incidence of covid-19 was observed (Figure 1). The proportion of patients with severe forms of covid-19 in the first wave was 5% and after the introduction of the P.1 variant this value almost doubled, quickly reaching values close to 10% between EW 6 and 9. The increase in the total number of covid-19 cases associated with an increase in the proportion of severe cases led to an abrupt increase in the number of patients admitted from the second half of February (Figure 1).

**Figure 1:**
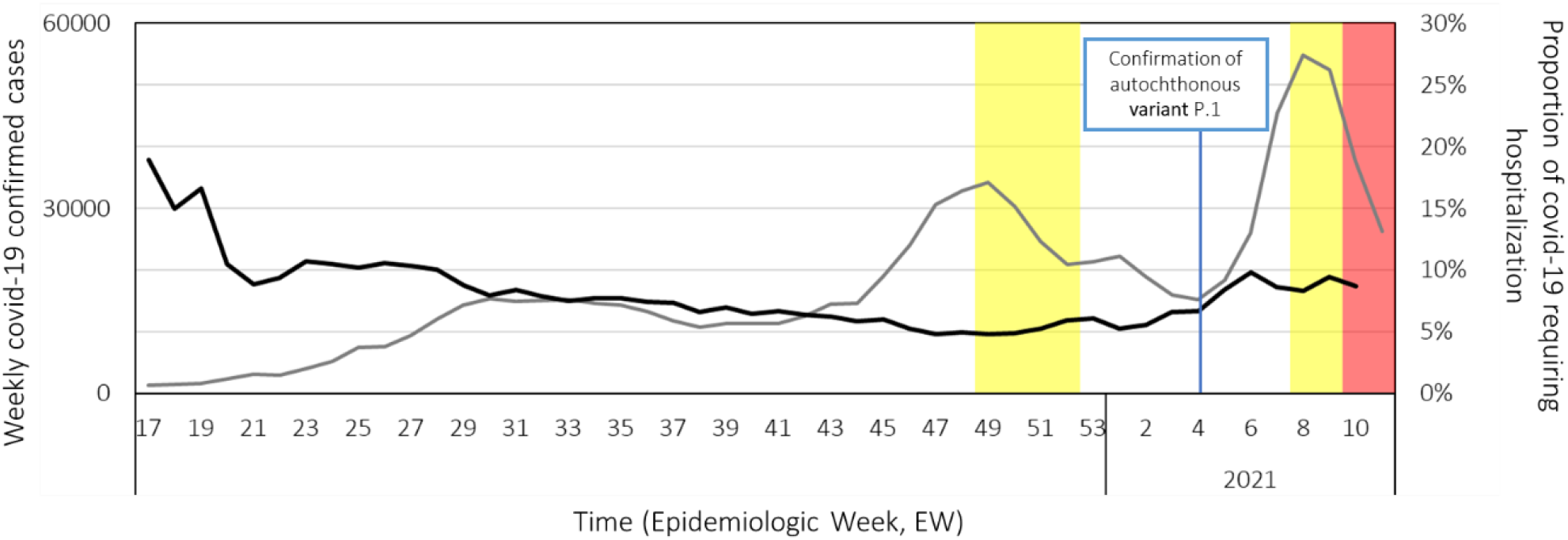
Grey line: number of new covid-19 cases (scale on the left); Black line: proportion of covid-19 cases requiring hospitalization by severe acute respiratory infection (SARI) cases among all covid-19 cases(scale on the rigth). Yellow area: time period when the bed ICU occupancy rate is between 80-100%; Red area: time period when the bed ICU occupancy rate is over 100%.

From the end of January, an increasing tendency in the hCFR overlaps with the begging of the local transmission of the P.1 variant. This increase occurred 2 weeks before the increase in the number of hospitalized patients and preceded by 4 weeks the exhaustion of the ICU beds, which occurred from March 3 (Figure 2).

**Figure 2:**
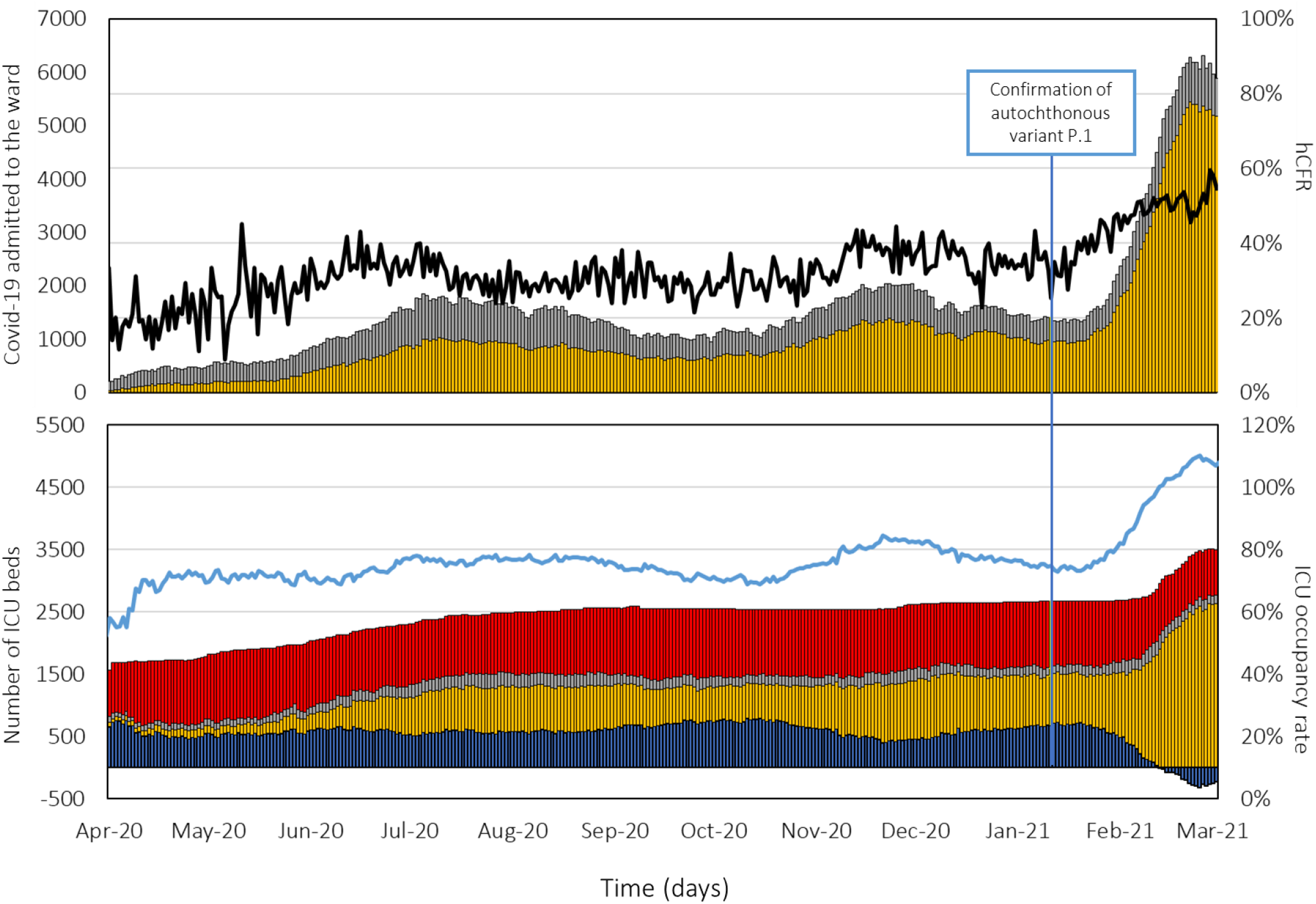
Above: number of beds occupied with covid-19 admitted to the ward (scale on the left) -yellow column confirmed cases, gray columns suspected cases, black line hospital case fatality rate (hCFR, scale on the rigth). Below: number of beds occupied with patients admitted to the ICU (scale on the left), yellow columns of covid-19 confirmed, gray columns suspected of covid-19, red columns other causes de admission, blue columns of beds available (negative numbers indicate that there is more demand than bed available), blue line ICU occupancy rate (scale on the right, values greater than 100% days when the health network had more demand than supply).

The case fatality rate also has increased in all age groups after the identification of the local transmission of the variant P.1 (figure 3). The age groups of 20 to 39 years old and of 40 to 59 years old presented a higher proportional increase in the second wave than in the first one (figure 3). The increase in the CFR begins to be noticed as of EW 4, that is, 6 weeks before the exhaustion of ICU vacancies in the state.

**Figure 3:**
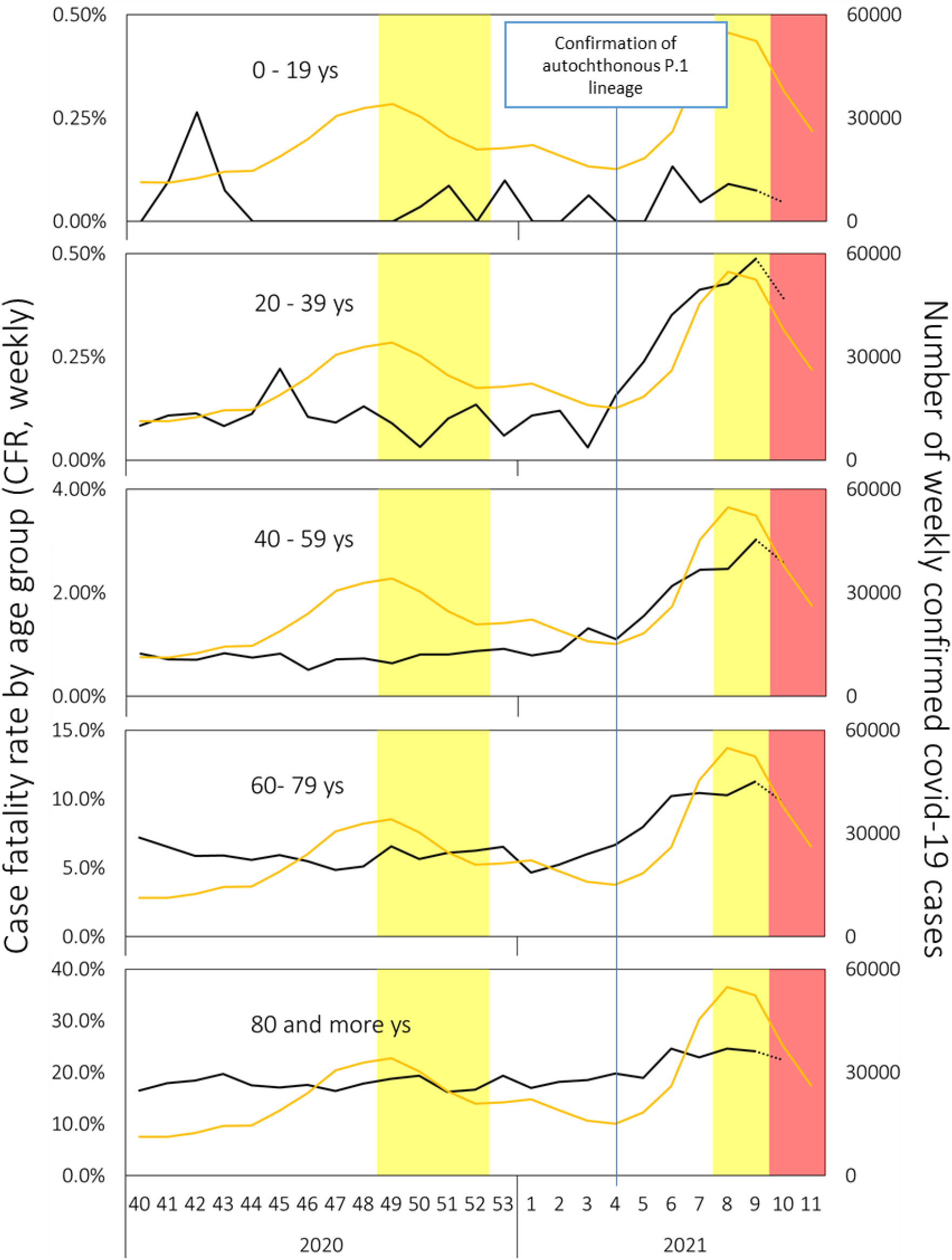
Black line: case fatality rate (CFR, %) of covid-19 confirmed cases according epidemiologic week of symptom onset in Rio Grande do Sul (Brazil) by age group(scale on the left). Orange line: number of weekly confirmed covid-19 cases (scale on the rigth). The yellow area corresponds to the period in which the ICUs were more than 80% full and less than 100% of the capacity, the red area corresponds to the period in which the ICUs were more than 100% of the capacity. Epidemiological week 10 is dotted because the data are not yet definitive.

With the exception of the group of people under 20 years old, a general increase in the proportion of severe cases in different age groups and sex was observed (table 2). However, this increase was prominent in the population between 20 and 59 years old and among patients without pre-existing risk conditions. The proportion of severe cases raised in the second wave in both sexes [female RR= 1.7 (95% CI = 1.64 −1.76; p <0.0001); male RR= 1.66 (1.61 - 1.72; p <0.0001)] (Table 2).The increase in the proportion of severe cases was greater in the 20- to 39-year-old group and in both sex than in the other age groups [female RR = 2.24 (95% CI = 1.99 - 2.52; p <0.0001); male RR = 2.56 (95% CI = 2.31 - 2.84; p <0.0001)].

**Table 2:**
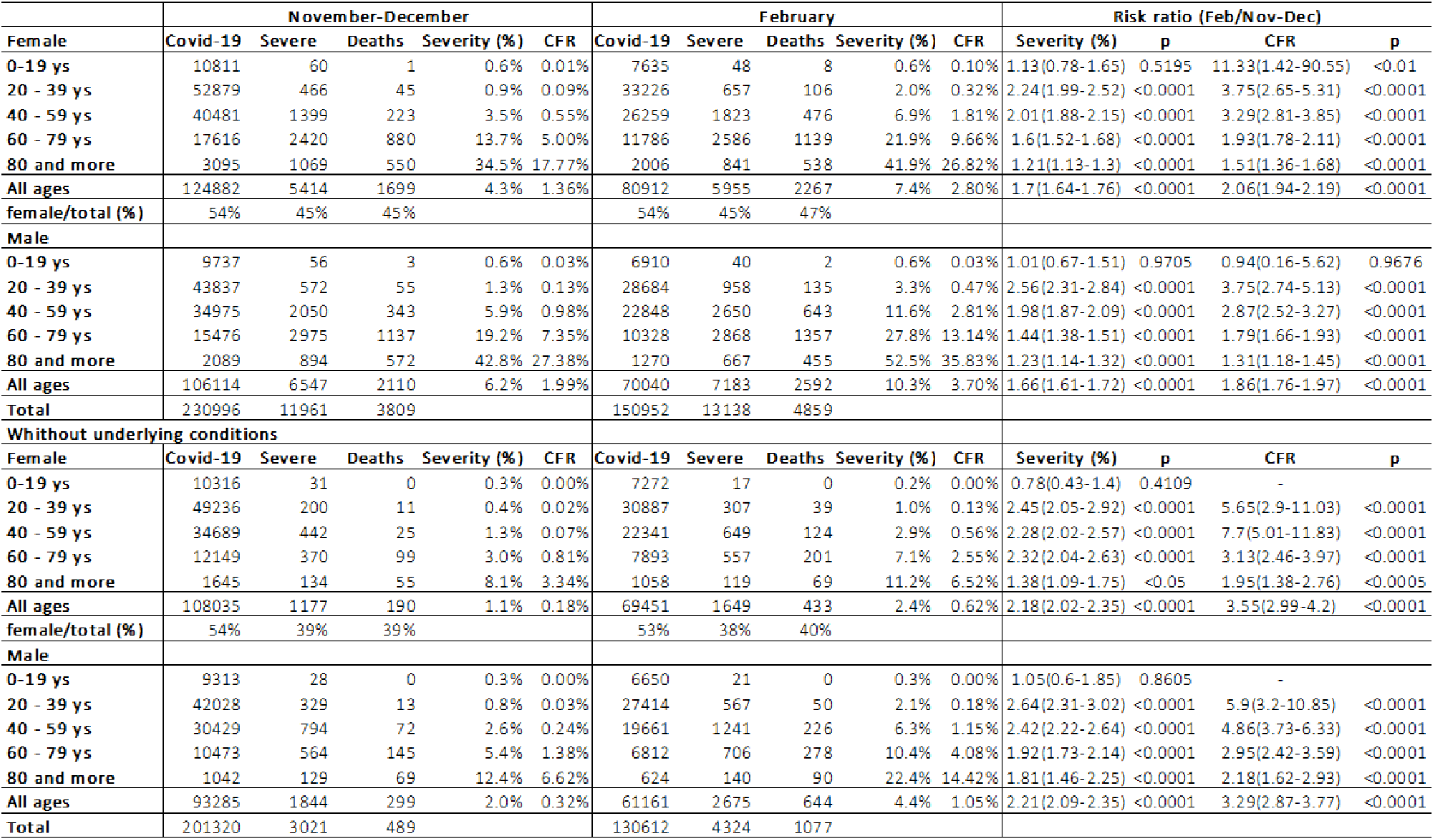
Cases, severe cases and deaths due to covid-19 and case fatality rate (CFR) during first (Nov-Dec/2020 and second waves (Feb/2021) by sex, age group and absence of underlying conditions in the Rio Grande do Sul state, Brazil..

The proportion of severe cases more than doubled among a population with no pre-existing diseases from the first to the second wave in both sexes (Table 2). In the age group between 20 and 39 years old, the increase in the proportion of severe cases was greater than in the other age groups [(Female RR = 2.45 (95% CI = 2.05 - 2.92; p<0.0001; Male RR = 2.64 (2.31 - 3.02; p <0.0001)].

Additionally, the CFR for covid-19 increased overall and in different age groups, in both sexes, when comparing the first and second waves. The increase in CFR occurred in a greatest intensity in the population between 20 and 59 years old and among patients without pre-existing risk conditions(table 2). Women in the 20 to 39 age group, with no pre-existing risk conditions, were at risk of death 5.65 times higher in February (95%CI = 2.9 - 11.03; p <0.0001) and in the age group of 40 and 59 years old, this risk was 7.7 times higher (95%CI = 5.01-11.83; p <0.0001).

The same profile was observed among men without previous diseases: the CFR was 5.9 (95%CI = 3.2 - 10.85; p <0.0001) higher in the second wave among adults between 20 and 39 years old and 4.86 higher (95%CI = 3.73 −6.33; p <0.0001) in February among men between 40 and 59 years old.

Among the population under 20 years of age, the CFR did not change, except for the female group. In this group, there was a significant increase (RR = 11.3 (1.42-90.55); p <0.01). However, this result should be analyzed with caution due to the large wide confidence interval and the small number of events in this age group.

## Discussion

Our results show that there was a general increase in the severity of cases and in the CFR for covid-19 in RS state after confirmation of the community transmission of variant P.1, and that this increase started before the overload of the health system and it was higher among young people without previous diseases. There was an increase in the proportion of patients under 60 years of age and of patients without pre-existing risk conditions between severe cases and deaths.

The increase in the proportion of hospitalized cases among cases of covid-19 in 2021 reversed a downward trend in the proportion of hospitalized patients that had occurred since the beginning of the pandemic, this fact preceded the increase in the total number of hospitalized patients. Thus, the increase in severity contributed to the overload of the local health system leading to the filling of the ICU beds. As we do not yet have specific treatment for covid-19 in the initial phase of the disease to prevent worsening, this increase in the proportion of severe cases cannot be attributed to problems in health care system. The fact that the increase in CFR and hCFR occurred simultaneously with the confirmation of the emergence of variant P.1 in the state and preceded by several weeks the overload of the health system reinforces the hypothesis that the risk of death among covid-19 patients increased regardless of overload of the hospital care system.

Although there was no shortage of beds in the wards during any analyzed period of the pandemic, a proportion of the cases with symptoms that started in late February may have had difficulty in hospitalization in the ICU in March and this may have contributed to the greatest increase in CFR and in hCFR. However, this does not explain why the difference was more striking in the population of young adults, without previous diseases, than between the elderly and the population in general. Vaccination also does not seem to be a plausible explanation for the risk changes between the age groups and in the population with pre-existing diseases, since at the end of February the vaccine was only being applied to health professionals, institutionalized elderly and over 80 years old. In this age group, until February 28, 48% (156,440 individuals) had recently received the first dose and only 0.7% (2,307 individuals) had received the second dose of the covid-19 vaccine (vacina.saude.rs.gov.br).

### Comparison with related studies

Our studies reinforce previous findings that indicate that a P.1 variant appears to be more lethal than the previous one, especially among young patients[2,4,13]. In a previous study in Amazonas, we found that in the subgroup of female patients, the difference in risk of death was greater than in the male population, in the present study we found similar results[2].

Studies in the United Kingdom found an increase in CFR in patients with a confirmed diagnosis of variant B.1.1.7 compared to the risk ratio of previous strains 1.64 (95% confidence interval 1.32 to 2.04)[14] and 1.61 (1.42-1.82) [15].

The increase in risk of death among variant B.1.1.7 patients in the United Kingdom calculated from dividing the CFR among B.1.1.7 patients by the CFR among non-B.1.1.7 patients was relatively homogeneous in the different age groups and in both the sexes. In females, it ranged from 1.46 in those over 85 years old to 1.59 in those under 35 years old. In males, it ranged from 1.47 in those over 85 years old to 1.55 in 35-54 years old[15].

Our study revealed a great variability in RR among the different groups by sex, age group and presence of pre-existing conditions. The heterogeneity observed between the age groups was greater when we analyzed the subgroup of the population without preexisting risk conditions where we found that the CFR in the female sex in the second wave was 1.95 times (95CI = 1.38-2.76) the CFR of the first wave in the population over 85 years old and was 7.7 times (95% CI = 5.01-11.83; p <0.0001) in the population between 40 and 59 years old. In the male population without previous diseases, the CFR in the second wave was 2.18 (95% CI 1.62-2.93) times the CFR of the first wave in the population over 85 years old and 5.9 (95% CI 3, 2-10,85; p <0, 0001) higher in the range between 20 and 39 years old. This heterogeneity occurs not only in the RR of the CFR, but also in the increase in the proportions of severe forms (RR of Severity, table 2), which excludes the impact of the overload of the health system, as we mentioned above.

### Implications of our findings

Brazil has one of the worst epidemiological situations for covid-19 in the world, both in the number of cases and in the number of deaths and their rates.[13] The situation has worsened greatly since the emergence of the P.1 variant, first in the Amazon region and then throughout the country. The presence of a large proportion of patients already infected in that region may have contributed to the selection of a strain with an immune escape capacity[16]. This phenomenon may also explain the emergence of a more transmissible strain, but not a more lethal one. A year has passed since the beginning of the SARS-CoV-2 pandemic and a major change in the epidemiological scenario appears to be the emergence of these new variants. Previous studies already exist that the P.1 variant is more transmissible and has the capacity to escape immunological.[3,8] The present study suggests that this variant leads to more severe and lethal results than previous strains. In addition, this variant was able to increase severity in specific groups that were previously more spared (women, youth and patients without pre-existing risk conditions). This set of characteristics should be an alert and reinforce the importance of better understanding the epidemiological, virological, pathophysiological, immunological and clinical aspects associated with this and other variants, therefore, an international effort must be organized to increase this knowledge.

### Strengths and limitations

One of the strengths of our study is its size, which includes all cases of covid-19 from the RS state officially reported to the Ministry of Health during a period when the P.1 variant was introduced in the state. Another strength is the fact that it is studying a period in which there was not yet a complete depletion of health resources in the studied place.

This study has some limitations, such as the use of secondary data, which can present problems in the quality, integrity and delays in the recorded data. Patients were not tested individually to see if it was a strain of SARS-CoV-2, for comparison of groups. We assume that as of February, the proportion of variant P.1 among patients with covid-19 was higher than in November and December, although it is quite likely, considering other studies in reference units in the state, this assumption is quite inaccurate.

The use of CFR and hCFR as severity indicators based on epidemiological surveillance data can lead to different types of bias, however, it remains a useful tool, especially in emergency situations in public health.[17–20]

This is an initial analysis in which a limited number of available variables have been assessed and additional individual risk factors have not been explored in depth. Despite these limitations, the results obtained in this study suggest a temporal association between the emergence of variant P.1 and severity indicators related to covid-19 measured by CFR and hCFR, in addition to a potential causal association between exposure to the new variant of SARS -CoV-2 (P.1). Therefore, subsequent specific studies must be carried out to evaluate these hypotheses.

### Conclusions

Our findings showed an increase in the proportion of young people and people without previous illnesses among severe cases and deaths in the state of RS after the identification of the local transmission of variant P.1 in the state. There was also an increase in the proportion of severe cases and in the CFR, in almost all subgroups analyzed, this increase was heterogeneous in different age groups and sex. As far as we know, these are the first evidences that the P.1 variant can disproportionately increase the risk of severity and deaths among population without pre-existing diseases, suggesting related changes in pathogenicity and virulence profiles. New studies still need to be done to confirm and deepen these findings.

## Data Availability

https://opendatasus.saude.gov.br/dataset/bd-srag-2021
https://ti.saude.rs.gov.br/covid19/

## Contributions of authors

André Ricardo Ribas Freitas: conceptualisation, data curation, formal analysis, investigation, methodology, project administration, validation, visualisation, writing original draft, and writing review & editing.

Daniele Rocha Queiróz Lemos: writing original draft, writing – review & editing Otto Albuquerque Beckedorff: data curation, have accessed verified the underlying data, writing – review & editing.

Luciano Pamplona de Góes Cavalcanti: writing – review & editing Andre M Siqueira: writing – review & editing

Regiane Cristina Santos de Mello writing – review & editing

Eliana Nogueira Castro de Barros: writing original draft, writing – review & editing

## Supplement

### Confirmed case of COVID-19 by laboratory criteria

- Detectable result for SARS-CoV-2 performed by the RT-PCR method, which detects in a sample of secretions from the airways (nose and throat) the SARS-CoV-2 virus, which causes COVID-19.
- Reagent result in an immunochromatographic or immunofluorescence antigen test that detects the SARS-CoV-2 virus.
- Reagent result in serological tests (rapid antibody tests, electrochemiluminescence, immunoenzymatic assay, among others), which identify antibodies produced in response to SARS-CoV-2 infection.

### Confirmed case of COVID-19 by clinical-epidemiological criteria

Case of SG or SRAG, without laboratory confirmation, with a history of close or home contact, in the 14 days prior to the appearance of signs and symptoms, with case confirmed laboratory for COVID-19.

### Confirmed case of COVID-19 by clinical-image criteria

Case of SG or SARS or death due to SARS that could not be confirmed by laboratory criteria and that presents tomographic changes indicative of SARS-CoV-2 infection.

### Confirmed case of COVID-19 by clinical criteria

Case of SG or SRAG associated with anosmia (olfactory dysfunction) or ageusia (gustatory dysfunction) without any previous cause and which could not be closed due to another confirmation criterion.

### Pre-existing conditions considered at risk

Obesity;

Myocardiopathies of different etiologies (heart failure, ischemic cardiomyopathy, etc.);

Arterial hypertension;

Cerebrovascular disease;

Severe or decompensated lung diseases (moderate / severe asthma, COPD);

Immunodepression and immunosuppression;

Chronic kidney disease in advanced stage (grades 3, 4 and 5);

Diabetes mellitus, according to clinical judgment;

Chromosomal diseases with a weakened immune status;

Malignant neoplasm (except non-melanotic skin cancer);

Hepatical cirrhosis;

Some hematological diseases (including sickle cell anemia and thalassemia);

Pregnancy

### The RECORD statement – checklist of items, extended from the STROBE statement, that should be reported in observational studies using routinely collected health data

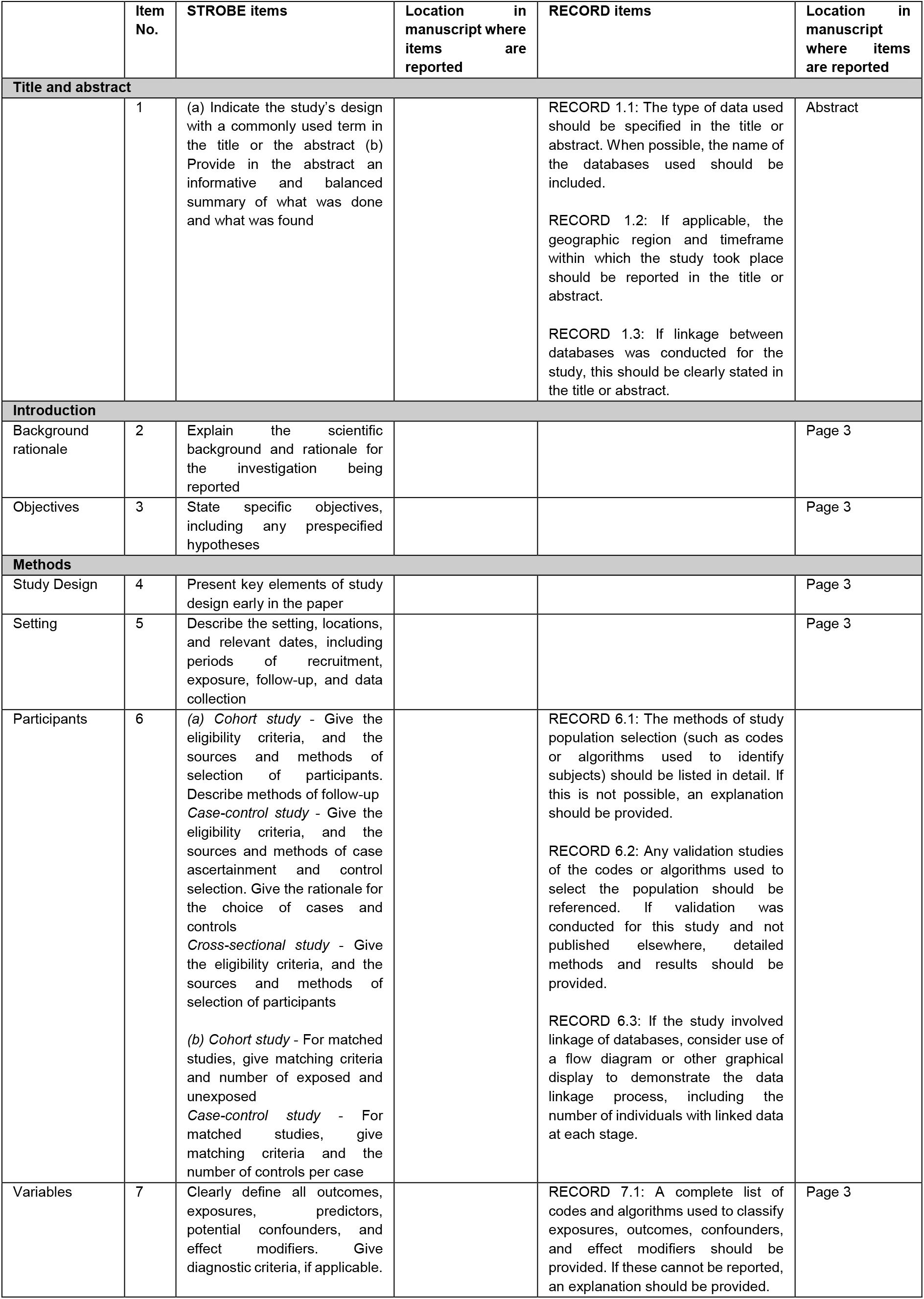
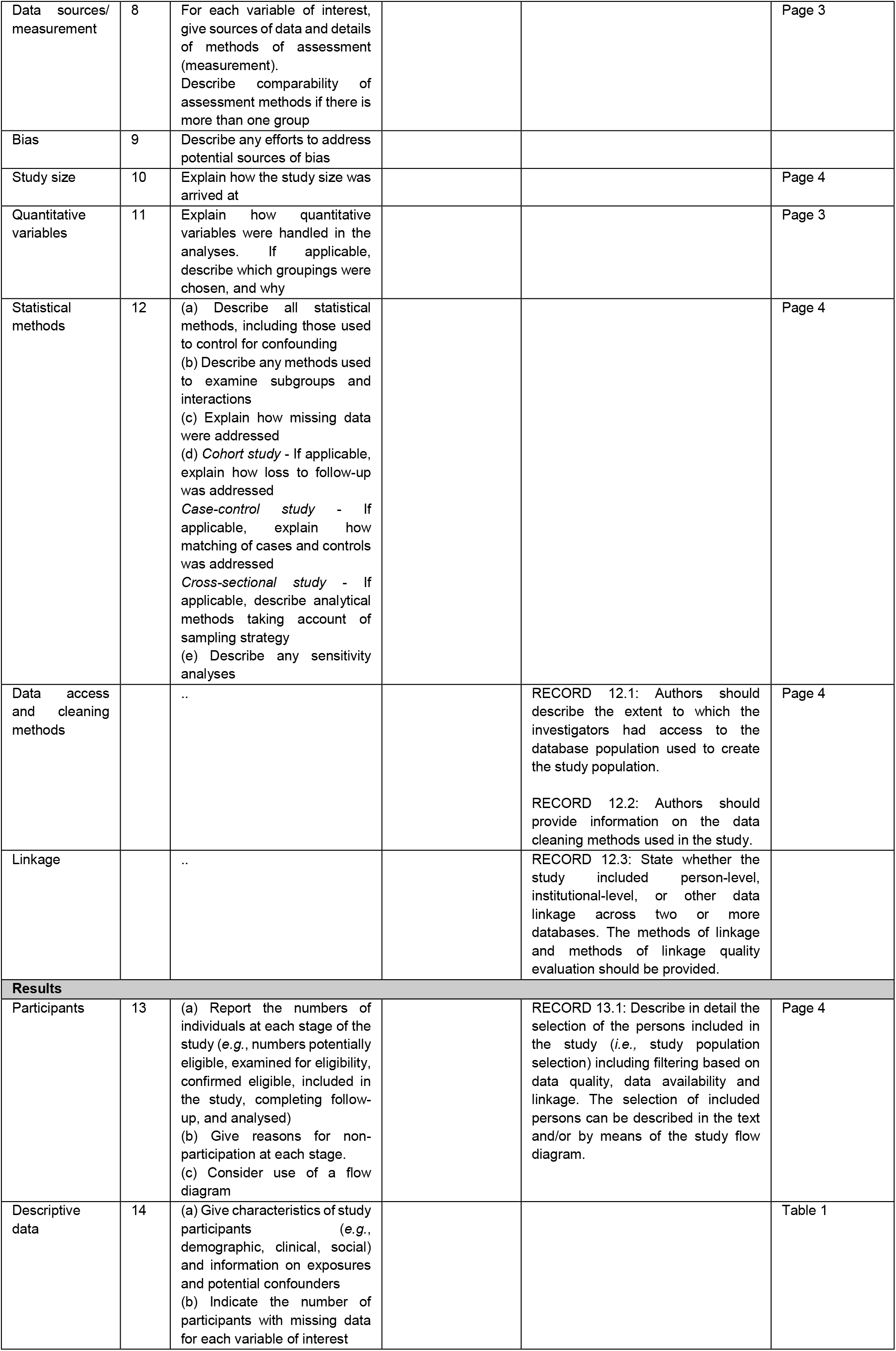
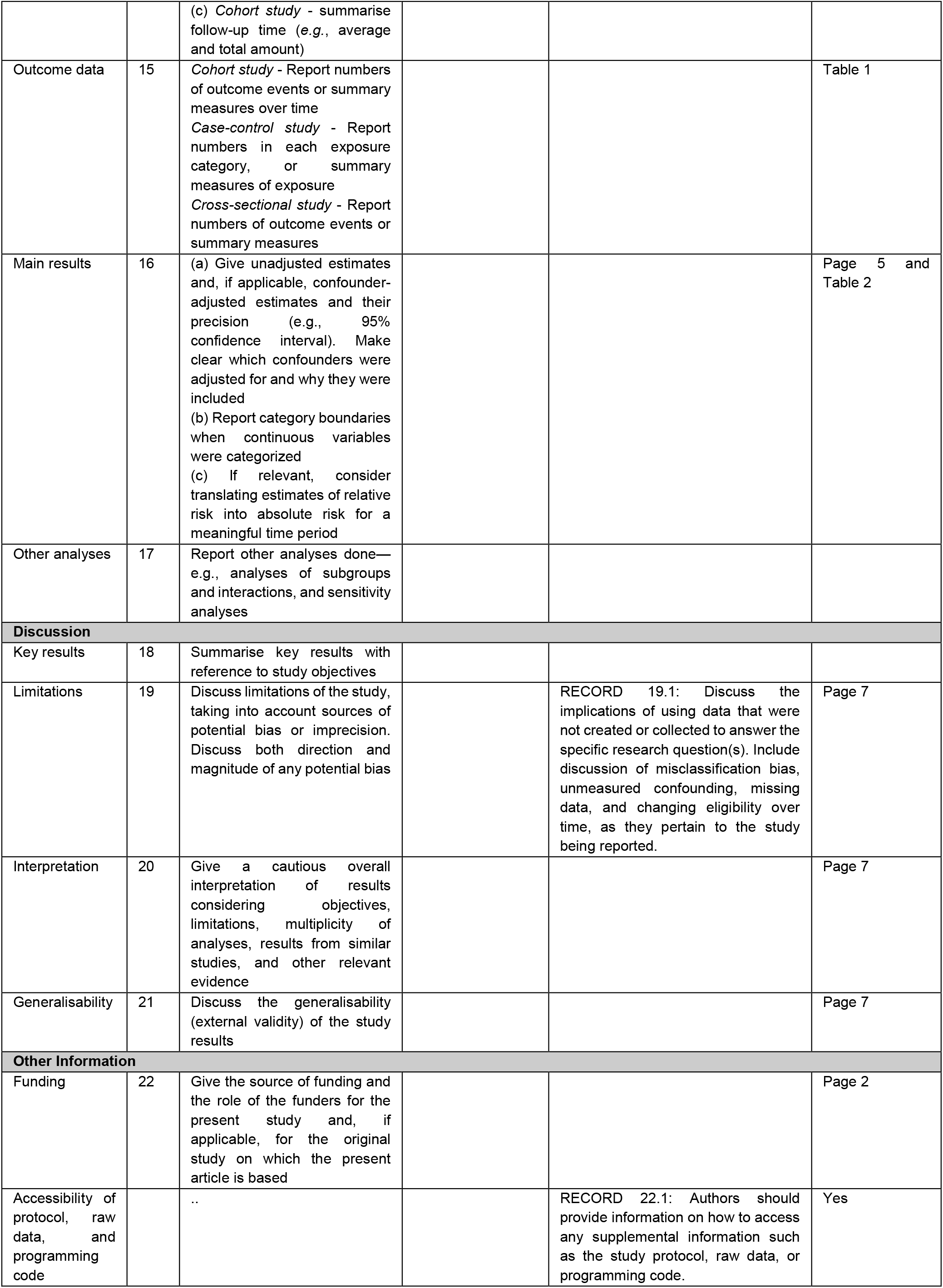

